# The Medical Action Ontology: A Tool for Annotating and Analyzing Treatments and Clinical Management of Human Disease

**DOI:** 10.1101/2023.07.13.23292612

**Authors:** Leigh C Carmody, Michael A Gargano, Sabrina Toro, Nicole A Vasilevsky, Margaret P Adam, Hannah Blau, Lauren E Chan, David Gomez-Andres, Rita Horvath, Megan L Kraus, Markus S Ladewig, David Lewis-Smith, Hanns Lochmüller, Nicolas A Matentzoglu, Monica C Munoz-Torres, Catharina Schuetz, Berthold Seitz, Morgan N Similuk, Teresa N Sparks, Timmy Strauss, Emilia M Swietlik, Rachel Thompson, Xingmin Aaron Zhang, Christopher J Mungall, Melissa A Haendel, Peter N Robinson

## Abstract

Navigating the vast landscape of clinical literature to find optimal treatments and management strategies can be a challenging task, especially for rare diseases. To address this task, we introduce the Medical Action Ontology (MAxO), the first ontology specifically designed to organize medical procedures, therapies, and interventions in a structured way. Currently, MAxO contains 1757 medical action terms added through a combination of manual and semi-automated processes. MAxO was developed with logical structures that make it compatible with several other ontologies within the Open Biological and Biomedical Ontologies (OBO) Foundry. These cover a wide range of biomedical domains, from human anatomy and investigations to the chemical and protein entities involved in biological processes.

We have created a database of over 16000 annotations that describe diagnostic modalities for specific phenotypic abnormalities as defined by the Human Phenotype Ontology (HPO). Additionally, 413 annotations are provided for medical actions for 189 rare diseases. We have developed a web application called POET (https://poet.jax.org/) for the community to use to contribute MAxO annotations.

MAxO provides a computational representation of treatments and other actions taken for the clinical management of patients. The development of MAxO is closely coupled to the Mondo Disease Ontology (Mondo) and the Human Phenotype Ontology (HPO) and expands the scope of our computational modeling of diseases and phenotypic features to include diagnostics and therapeutic actions. MAxO is available under the open-source CC-BY 4.0 license (https://github.com/monarch-initiative/MAxO).

## Introduction

Numerous computational resources exist to annotate and analyze pharmaceutical data, including DrugCentral, DrugBank, and the Pharmacogenomics Knowledge Base (PharmGKB).^1–3^ However, clinical management includes many other measures that collectively we refer to as medical actions. For instance, clinicians may prescribe a certain dietary intervention such as coenzyme Q10 supplementation for individuals with primary coenzyme Q10 deficiencies, or recommend the avoidance of a ketogenic diet, which although helpful for some individuals with epilepsy is contraindicated for individuals with certain diseases.^4^ Other medical actions include surgical procedures, ablations, treatment with biologics, behavioral and cognitive interventions, deep brain stimulation, and many others.

In medical practice, it is commonplace that the treatment of a condition can depend both on the specific subtype of a condition as well as the underlying disease, if any, that led to the condition. For instance, occasional tension-type headaches are easily treated with non-prescription medications such as aspirin or acetaminophen, but specific categories of headaches such as migraine may require other medications, and other subtypes such as primary exertional headaches, may require modification or avoidance of exercise regimens.^5^ In addition to this, rare diseases may present with headaches that require specific diagnostic or therapeutic measures that would not typically be indicated in other settings. For instance, individuals with Marfan syndrome may present with postural headaches related to intracranial hypotension due to dural ectasia, which can be diagnosed by spinal magnetic resonance imaging;^6^ migraine is a common manifestation for individuals with cerebral autosomal dominant arteriopathy with subcortical infarcts and leukoencephalopathy (CADASIL), and these individuals may respond more poorly to beta-blocker treatment than the general population of individuals with migraine;^7^ headache may occur as a manifestation of ornithine transcarbamylase deficiency, but clinical management should focus on acute and chronic reduction of elevated ammonia levels in the blood circulation.^8^ In addition to this, an increasing number of rare disease-specific treatments and clinical management options are being developed including gene therapy, dietary regimens, and biologics.^9^ An increasing number of therapeutics for people with rare diseases (RDs) have been developed, although it is estimated that effective treatments are available for less than 5% of the more than 10,000 rare diseases believed to affect humans.^10^

Although RDs are highly heterogeneous in etiology, clinical presentation, and clinical management strategies, they share commonalities related to their very rarity: a lack of knowledge and scarcity of expertise.^11^ The scarcity of expertise about RDs among many physicians is probably one of the reasons for the often-bemoaned diagnostic odyssey that many individuals with RDs experience: it has been estimated that the average time for accurate diagnosis of an RD is 4-5 years, with the diagnosis being delayed over a decade in some cases.^12^ The same factors that delay diagnosis also delay proper disease management even after a diagnosis has been made; often available therapies are identified only after the therapeutic window has passed.

In this work, we present the Medical Action Ontology (MAxO), a comprehensive open-source computational representation of medical diagnostics, preventions, procedures, interventions, and therapies; MAxO is designed to apply to any patient at any age or stage of life and experiencing any kind of disorder. MAxO is the first terminology to systematically cover medical actions. MAxO is an interoperable OBO foundry ontology with a rich representation of terms by providing definitions and synonyms to take into account the broader context of each term. There are currently over 1757 terms in MAxO. Moreover, as part of the Monarch Initiative^13^ suite of ontologies, MAxO’s medical actions have also been used to annotate disease and disease manifestations. While the initial focus and use case for MAxO is RD, MAxO is designed to be able to address all clinical cases and diseases. This manuscript introduces MAxO, describes its intended use cases, and presents POET, a web-based curation tool designed to facilitate community participation in the future growth of MAxO and MAxO disease annotations.

## Materials and Methods

### Ontology Curation and Governance

MAxO was created and maintained using standard ontology engineering practices and executable workflows provided by the Ontology Development Kit (ODK).^14^ MAxO files are publicly available on GitHub (https://github.com/monarch-initiative/MAxO). The ODK provides functionality to ensure consistent release versioning, managing dependencies on other ontologies, and quality testing. Whenever a change to the ontology is proposed, a battery of tests is run to ensure that all terms have one and only one label and definition, are logically consistent, and are classified in the right branch of the ontology. MAxO is edited using Protégé (currently, version 5.6.1).^15^ To maintain ontological consistency throughout MAxO we employ ontology design patterns for most of the logical axiomatization, described in the next section. MAxO is an OBO Foundry (OBO) ontology^16^ and adheres to OBO best practices including providing an open license (CC-by 4.0 license), distributing in a common format, and maintaining interoperability with external ontologies. MAxO is iteratively developed and regular releases are made available on GitHub (https://github.com/monarch-initiative/MAxO/releases). We welcome community requests for new terms or changes to current terms (such as reclassification, revision to definitions, addition or removal of synonyms, etc.) via tickets on our GitHub issue tracker (https://github.com/monarch-initiative/MAxO/issues).

### Using Design Patterns and Templates for consistent creation of terms

Terms included in MAxO must qualify as a medical action. We define ‘medical action’ broadly as a clinical procedure, a diagnostic investigation, a therapy, an intervention, or a recommendation required in the care of patients. For every medical action we include, we define a label that reflects common practice, a human-readable definition, and provenance metadata (such as who contributed the term). In many cases, synonyms (exact, narrow, or broad) are used to further explain the meaning of the term as well as be used in text mining algorithms. Sometimes there are other controlled vocabularies (e.g. NCIT) or ontology terms that have the same or similar meaning. In these cases, we included suitable cross-references. Many medical actions included in MAxO were identified during the curation of *GeneReviews^®^*, which currently comprises 864 chapters on single genes or diseases or overviews of disease groups.^17^ Many others were identified during Human Phenotype Ontology (HPO)^18^ annotation curation. Other terms were identified from PubMed, Google Search, OMIM, and other combinations thereof.

Dead Simple Ontology Design patterns (DOSDP) provide a framework for the consistent generation of terms according to well-defined templates, both for metadata (synonyms and labels) and logical axiomatization.^19^ DOSDPs allow us to programmatically generate new terms that afford internal logical consistency across medical actions in MAxO but also consistent logical links to other ontologies such as FoodOn and ChEBI. For example, the “nutrition supplementation” pattern allows us to define new supplementation terms such as “folic acid supplementation” by simply pointing to the concept of “folic acid” in ChEBI. DOSDP tools then generate a set of suitable definitions, synonyms, and axioms. This DOSDP pattern together with the same-named TSV file generates 58 nutrition supplementation terms. For use cases that do not require logical patterns (such as the documentation of deprecated terms and their replacements or mappings), we make use of ROBOT templates^20^ instead of DOSDP, which do not provide any functionality to automatically generate consistent axioms, but are simpler to use. Specific information about how the templates were set up can be found on MAxO’s wiki page (https://github.com/monarch-initiative/MAxO/wiki/DOSDP-pattern-creation-and-edits) or in tutorials here (https://oboacademy.github.io/obook/tutorial/dosdp-template/).

### Curation of diagnostic procedures for phenotypic abnormalities

The diagnostic branch of MAxO was used to annotate diagnostics to phenotypes using Human Phenotype Ontology (HPO) terms. These annotations are disease agnostic. For instance, ‘Prolonged QTc interval’ (HP:0005184) can be diagnosed by *electrocardiography* (MAXO:0000900), regardless of the underlying disease. Suggestions for additional diagnostic annotations are welcome via the HPO GitHub issue tracker (https://github.com/obophenotype/human-phenotype-ontology/issues). Disease-specific diagnostic annotations will be curated as discussed below.

### Community curation of Medical Actions for Rare Disease with POET

POET (https://poet.jax.org/) is a modern full-stack application to generate annotations for disease and associated ontologies. POET provides a concise interface for scientific curators to submit MAxO annotations (see below for details on annotation structure). Using phenotypes annotated to rare diseases (https://hpo.jax.org/app/data/annotations), MAxO terms can be annotated directly to disease or to the disease manifestations using this web tool. Full documentation is available at https://monarch-initiative.github.io/maxo-annotations/.

## Results

Each term in MAxO describes a medical action such as *bronchoscopy* (MAXO:0001183). We define medical action broadly as any medical procedure, intervention, therapy, and or measurement undertaken for clinical management. The structure of the MAxO, which allows a term to have multiple parent terms, enables different aspects of phenotypic abnormalities to be explored (Figure 1).

**Figure 1.**
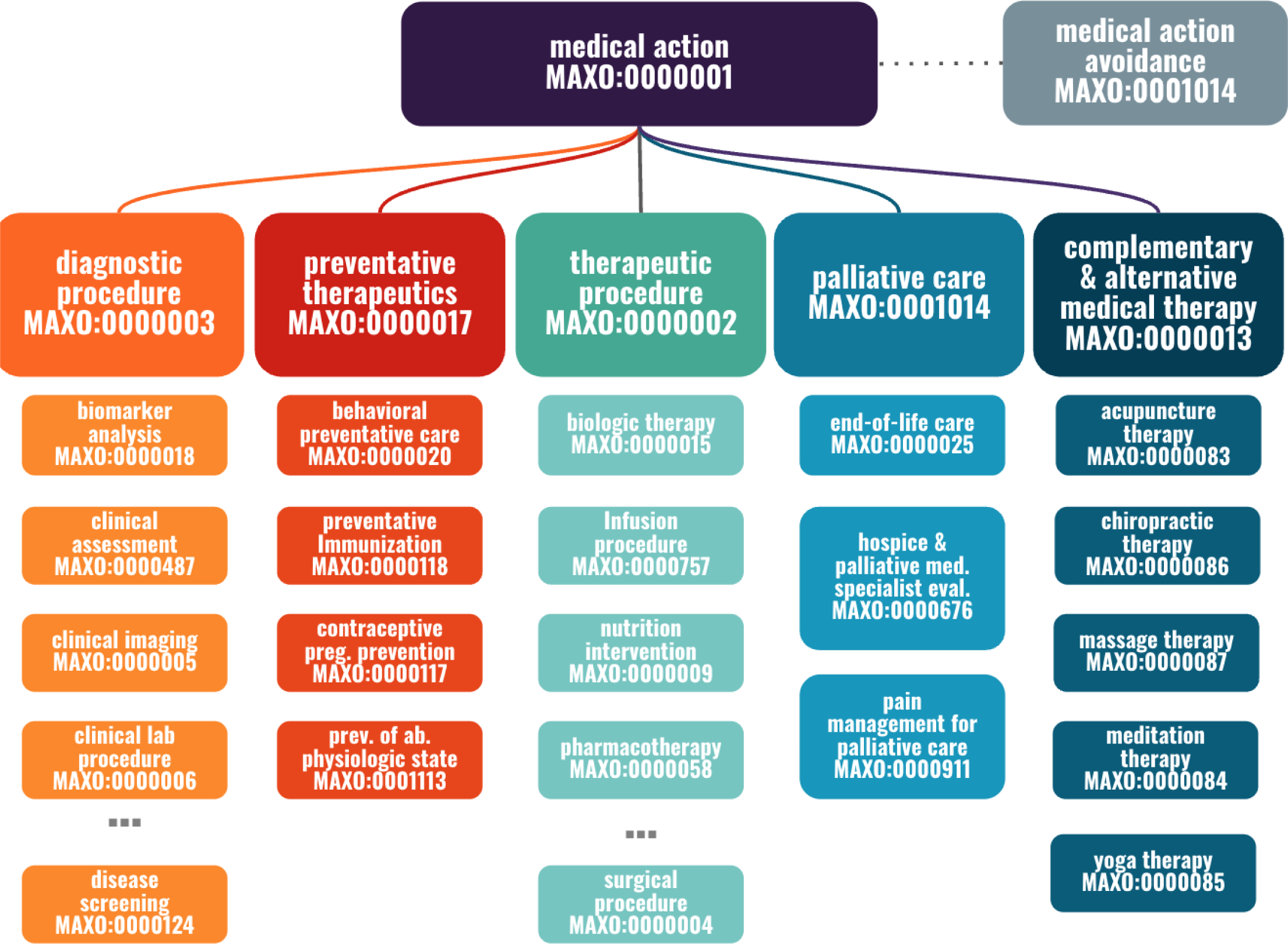
Structure of the Medical Action Ontology (MAxO). (A) The upper level of MAxO comprises six categories of medical actions: *diagnostic procedure*, *preventative therapy*, *therapeutic procedure*, *medical action avoidance*, *palliative care*, and c*omplementary & alternative medical therapy*. The subhierarchy *therapeutic procedure* (MAXO:0000002) contains the largest number of terms. ‘…’ indicates additional terms are present in that hierarchy.

There are currently 1757 terms with 4695 synonyms (as of the June 2023 release) and 6 upper-level terms that encompass all of MAxO. The diagnostic and therapeutic procedure branches currently contain the largest number of terms. The *therapeutic procedure* (MAXO:0000002) branch is where the majority of the terms reside, including *cognitive and behavioral interventions*, *hydrotherapies*, *nutrition interventions*, *pharmacotherapies, physical therapies*, *radiation therapy*, *supportive care*, and *surgeries*. Some *diagnostic procedures* are also present in the *therapeutic procedure* branch, as some techniques can be used both as diagnostics and therapeutics. The *diagnostic procedure* (MAXO:0000003) branch is the second largest branch of MAxO and currently contains 774 terms.

Additional high-level branches are *complementary and alternative medical therapy* (MAXO:0000013), *palliative care* (MAXO:0000021), *preventative therapeutics* (MAXO:0000017), and *medical action avoidance* (MAXO:0001014) (Figure 1) to address different aspects of clinical care. Complementary and alternative medical therapy are actions like *yoga therapy* (MAXO:0000085) and *acupuncture therapy* (MAXO:0000083) that are not part of the standard care, but some professionals may prescribe it for rare diseases such as Sickle Cell Anemia and Ehlers-Danlos Syndrome.^21,22^ *Preventative therapeutics* (MAXO:0000017) are defined as medical actions that prevent the occurrence of a disease or halt a disease and avert resulting complications after its onset.^23^ Some examples are vaccinations or smoking prevention. Recommendations can be to avoid or prevent an abnormal physiologic state, like obesity (*obesity prevention recommendation*; MAXO:0000047) or dehydration recommendation (*dehydration prevention recommendation*; MAXO:0000045). In Carnitine-Acylcarnitine Translocase Deficiency, it is recommended to avoid catabolic illness, including the recommendation to prevent fevers (*fever prevention recommendation;* MAXO:0001116).^24^ We additionally developed a branch to accommodate recommendations to avoid specific medical actions (*medical action avoidance;* MAXO:0001014). For instance, the term *avoid CT scan* (MAXO:0010321) applies to Nijmegen Breakage Syndrome, because it is recommended that affected individuals avoid unnecessary exposure to ionizing radiation, including CT scans.^25^. We note these annotations are not intended to convey medical recommendations but rather to report descriptions of clinical management in the literature. Clinicians should assess information in light of the clinical picture of each case and make informed risk-benefit assessments. Each term consists of an identifier, a preferred label, a definition, and information about its place in the ontology (subclass relation). Most terms have a list of synonyms and a computable definition (Table 1).

**Table 1.** Anatomy of a MAxO Term. Each MAxO term is created with a unique identifier (in this case, MAXO:0010012), a label, synonyms if available, a logical definition that specifies how the term related to terms from external ontologies and to other terms in MAxO, subclass relations, and information about the creator of the term and the creation date (not shown here).

|  |  |
| --- | --- |
| **id** | MAXO:0010012 |
| **label** | *coenzyme Q10 supplementation* |
| **definition** | Addition of coenzyme Q10 to the diet. |
| **synonyms** | coenzyme Q10 supplemental intake<br>supplementation of coenzyme Q10<br>dietary supplementation of coenzyme Q10 |
| **OWL logical definition** | nutritional supplementation<br>and |
|  | has input some coenzyme Q10 (CHEBI:46245) |
| **Subclass of** | Nutritional supplementation |
| **Created by** | http://orcid.org/0000-0001-7941-2961 |

### Logical structure of MAxO

Nearly half of the terms in MAxO were created using DOSDP templates (Methods; Figure 2). These are logical templates that enable the standardized creation of MAxO terms that are logically linked to other ontologies. Of the 1757 MAXO terms, 941 are computationally defined in this way with reference to 633 distinct terms from the Relation Ontology (RO),^26^ Uber-anatomy Ontology (Uberon),^27^ Gene Ontology (GO),^28^ Food Ontology (FoodOn),^29^ Chemical Entities of Biological Interest Ontology (ChEBI),^30^ HPO, Neurobehavior Ontology (NBO), and Basic Formal Ontology (BFO).^31^ Thus, MAxO is built from the start to link to many other sources of knowledge. This interlinkage offers the advantage that we can use OWL semantic reasoning to automatically infer the hierarchical placement of MAxO terms according to the corresponding structure of the source ontologies. For example, if a new term *glutamine supplementation*, associated with *glutamine*, is added to MAXO, it will be automatically classified as a kind of *amino acid supplementation*, because *glutamine* is a kind of *amino acid* in CHEBI.

**Figure 2.**
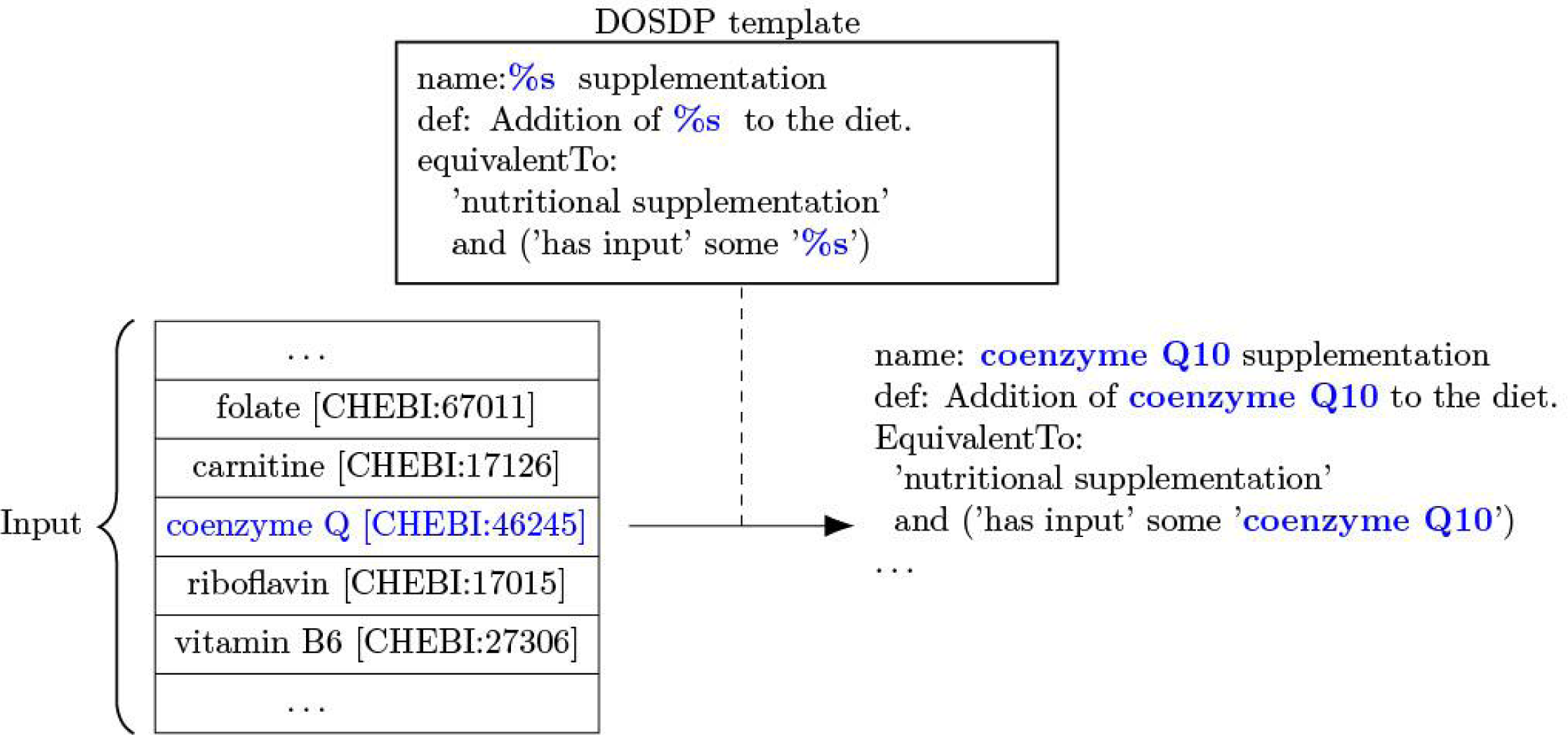
Templated MAxO term creation with DOSPD. New terms are created by specifying a list of terms from an ontology such as the Chemical Entities of Biological Interest (ChEBI) and using the DOSDP to create the MAxO terms according to a template that specifies the structure of each component of the term to be created. In this example, an excerpt of the template is shown that specifies how to create the term name, definition, and OWL class definition. DOSDP creates a new MAxO term for each item in the list. In the example, coenzyme Q10 is shown. The template “%s supplementation” specifies that the name of item in the list (“coenzyme Q10”) should be substituted to create the final name (“coenzyme Q10 supplementation”). The remaining, unshown term components are created analogously.

DOSDPs are useful to create a collection of related terms in a consistent fashion. For instance, MAxO currently contains 58 terms that descend from *nutritional supplementation* (MAXO:0000106). *Nutritional supplementation* is the administration of concentrated sources of nutrients or other substances with a nutritional or physiological effect that supplement the normal diet.^32^ The term *coenzyme Q10 supplementation* (MAXO:0010012) represents a treatment of diseases such as rare disease type 4 primary coenzyme Q10 deficiency (MONDO:0018151).^33^ The DOSDP template includes variable slots to define the term. For instance, the label *coenzyme Q10 supplementation* is generated by the template “%s supplementation” together with the term from ChEBI^30^ *coenzyme Q* (CHEBI:46245). (Figure 2) MAxO currently has 24 different DOSDP patterns importing terms from neurobehavior ontology (NBO), CHEBI, Food Ontology (FoodOn), human phenotype ontology (HPO), Uber-anatomy ontology (UBERON), and protein ontology (PRO) as well as using relationship ontology (RO) for relationships. Fourteen of these patterns heavily rely on UBERON for the anatomy ‘location’ of a procedure. For example, ‘mri_by_location’ DOSDP pattern uses the *medical magnetic resonance imaging procedure* (MAXO:0000424) and the UBERON anatomy term UBERON:0000916 to make terms such as *MRI of the abdomen* (MAXO:0000426). All these patterns are modular and customizable to be able to add terms quickly and/or add very specified curation.

### MAxO Therapeutic Annotations

The MAxO project currently includes 413 annotations to 189 RDs. The annotations use 161 unique MAXO terms to 212 unique HPO terms. Each annotation identifies a medical action and its relationship to a disease or disease manifestation. The MAxO term’s relation to the disease or disease manifestation is specified by one of five relations (Table 2). Some medical actions target the disease itself; for instance, *fava bean intake avoidance* (MAXO:0010051) can in principle prevent all clinical manifestations of *G6PD deficiency* (MONDO:0005775).^34^ In other cases, treatment is specific to a subset of disease manifestations. For instance, *removal of the lens* (MAXO:0001220) can be used to treat lens dislocation in *Marfan syndrome* (MONDO:0007947),^35^ but this measure does not treat other manifestations of Marfan syndrome such as aortic root dilatation.

**Table 2.**
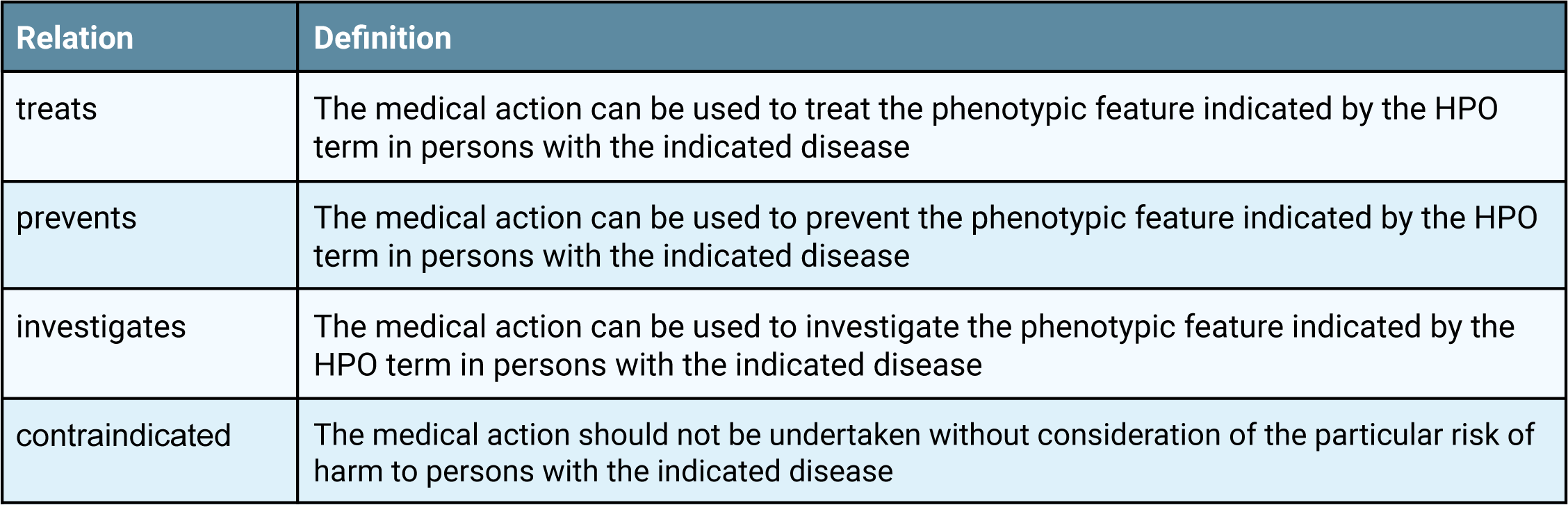
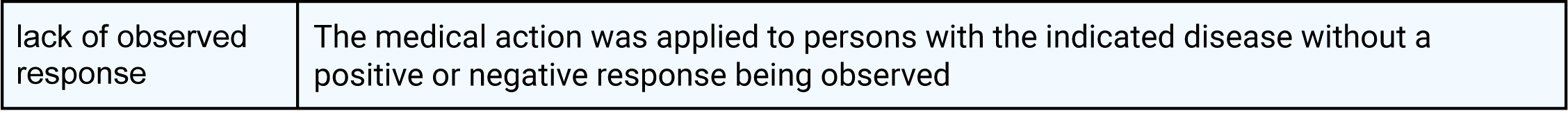
Definitions of MAxO Annotation Relations.

| Relation | Definition |
| --- | --- |
| treats | The medical action can be used to treat the phenotypic feature indicated by the HPO term in persons with the indicated disease |
| prevents | The medical action can be used to prevent the phenotypic feature indicated by the HPO term in persons with the indicated disease |
| investigates | The medical action can be used to investigate the phenotypic feature indicated by the HPO term in persons with the indicated disease |
| contraindicated | The medical action should not be undertaken without consideration of the particular risk of harm to persons with the indicated disease |
| lack of observed response | The medical action was applied to persons with the indicated disease without a positive or negative response being observed |

Therefore, the MAxO annotation model includes disease ID and label using the MONDO ontology ^13,36^, the MAxO term id and label, and an HPO term that represents the target of the treatment. If the treatment targets the disease itself rather than specific manifestations of the disease, this is indicated by the corresponding MONDO id. Finally, two fields for an extension ID and label can be used to specify additional information about a treatment. For instance, in the example, the MAxO term serotonin-norepinephrine reuptake inhibitor agent therapy is further specified with the ChEBI term for fluoxetine, which is a specific type of serotonin-norepinephrine reuptake inhibitor (Table 3). Lastly, an additional field is available for comments. In the example provided in Table 2, fluoxetine, a treatment commonly used for one type of congenital myasthenic syndrome (slow-channel congenital myasthenic syndrome, CMS 1A/2A/3A/4A), may worsen symptoms of fatigable weakness if administered to patients with a different type of congenital myasthenic syndrome caused by defects in the same gene, fast-channel CMS (CMS 1B/2B/4B). Fluoxetine is a common treatment for depression that may mistakenly be administered to patients affected by fast-channel CMS without understanding the need for its avoidance. The two MAxO entries listed in Table 2 are linked to the specific CMS subtypes involved and can thus indicate the precise diseases for which fluoxetine is an appropriate treatment (example 1) and in which it should be avoided, i.e., the treatment is contraindicated (Table 3).

**Table 3.**
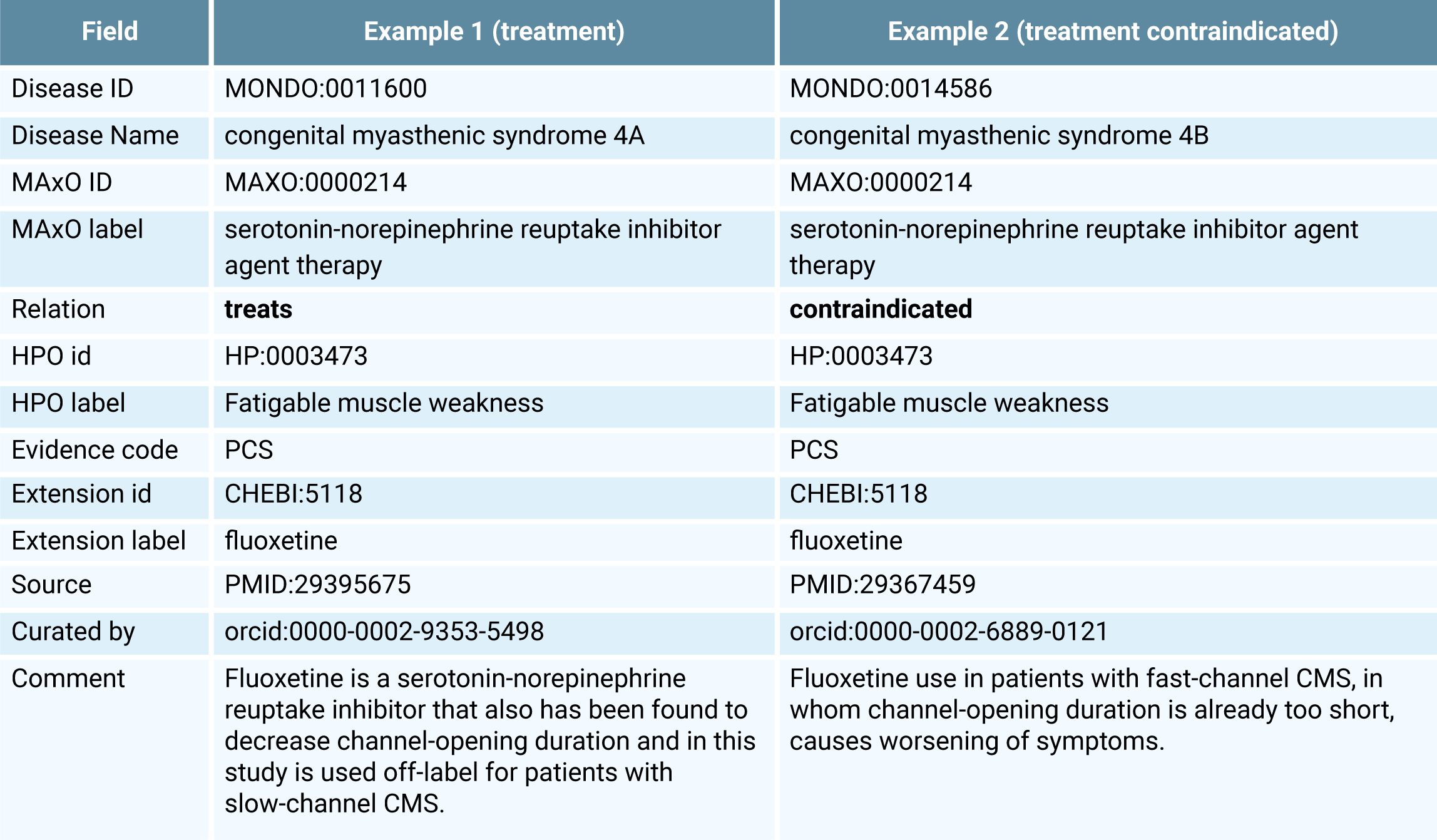
Annotation format for linking MAxO terms to diseases. Annotations are shown for *congenital myasthenic syndrome 4A* (MONDO:0011600) and *congenital myasthenic syndrome 4B (*MONDO:0014586). Both have the same disease annotations, but medical action *serotonin-norepinephrine reuptake inhibitor agent therapy* (MAXO:0000214) have different relations where it is a recommended treatment (**treats**) 4A, but it is contraindicated (**contraindicated**) in 4B. The evidence code PCS corresponds to ECO:0006016 (author statement from published clinical study).

## MAxO diagnostic annotations

With a few exceptions for special cases, HPO terms do not specify the diagnostic tools by which the features described by the terms can be ascertained. For instance, Agenesis of corpus callosum (HP:0001274) can be ascertained by methods including fetal ultrasound and magnetic resonance tomography imaging of the brain.^37^ We envision that it will be useful to specify which diagnostic modalities can be used to diagnose or exclude individual phenotypic abnormalities (i.e., HPO terms). We have therefore introduced a new relationship into the HPO, is_observable_through, and use it to annotate HPO terms with terms from the MAxO *diagnostic procedure* (MAXO:0000003) subontology. Currently, there are 18,505 diagnostic annotations comprising 16,389 unique HPO terms and 259 unique MAxO terms. Of these, 2068 specify whether prenatal (fetal) phenotypic abnormalities are ascertainable by prenatal sonography or magnetic resonance imaging (**Figure 3**).

**Figure 3.**
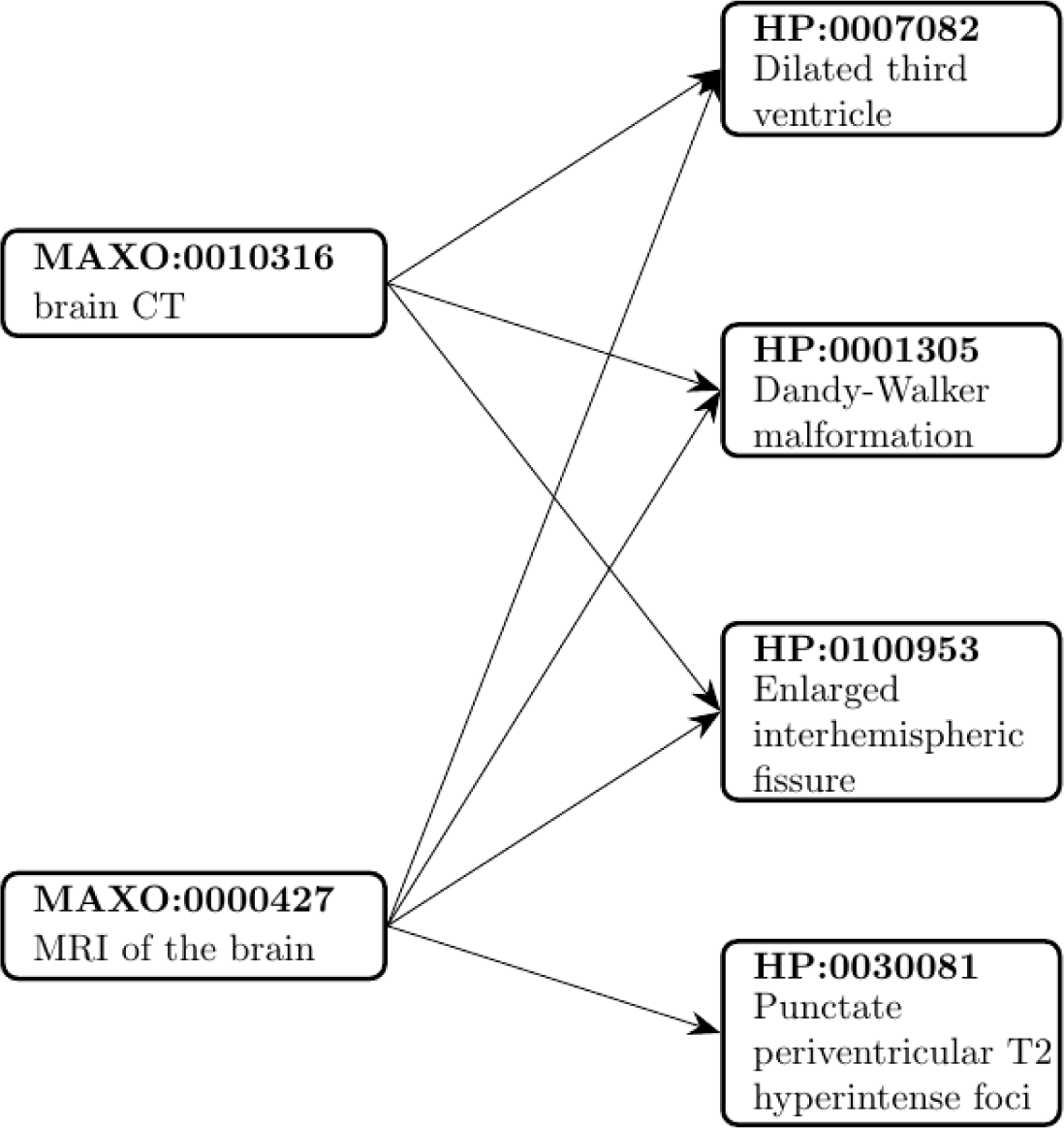
MAxO diagnostic annotations. In this example, the MAxO term brain CT (MAXO:0010316) annotates 108 HP terms (three of which are shown here), and MRI of the brain (MAXO:0000427) annotates 445 HP terms (four of which are shown here). Note that T2 hyperintense foci can be observed by MRI but not CT because T2 refers to a specific form of magnetic resonance imaging weighting (the same lesion may be apparent on CT but not by T2 intensity). A total of 18,493 diagnostic annotations are available in the June 2023 MAxO release.

## POET

POET (*Phenotype Ontology Engineering Tool*) is a web-based application that is designed to generate data for disease and associated ontologies. Built with the latest technologies (Spring Boot, Auth0, and Angular) and designed with scientific curators in mind, POET provides a concise interface for quickly and securely annotating phenotypes to rare diseases (https://hpo.jax.org/app/data/annotations) using MAxO terms. All annotations are seamlessly downloaded with the rolling ontology releases, ensuring efficient and accurate data management. Technical users alike can benefit from POET’s modern, user-friendly interface.

All annotations require a reference (PMID). Moreover, only previously disease-associated phenotypes will be able to be annotated with a medical action. That limits the unverified expansion of disease-associated phenotypes. In addition, annotations require a relationship to the phenotype (i.e., treats, investigates, contraindicated, lack-of-observed-response, or prevents), and evidence type (i.e., traceable author statement, clinical study, or inferred electronic annotation). These fields create the context for each annotation. Optional fields include an ontology extension and a comment section. Ontology extensions are used for pharmacotherapy terms where a specific CHEBI/drug term is indicated (Figure 4).

**Figure 4.**
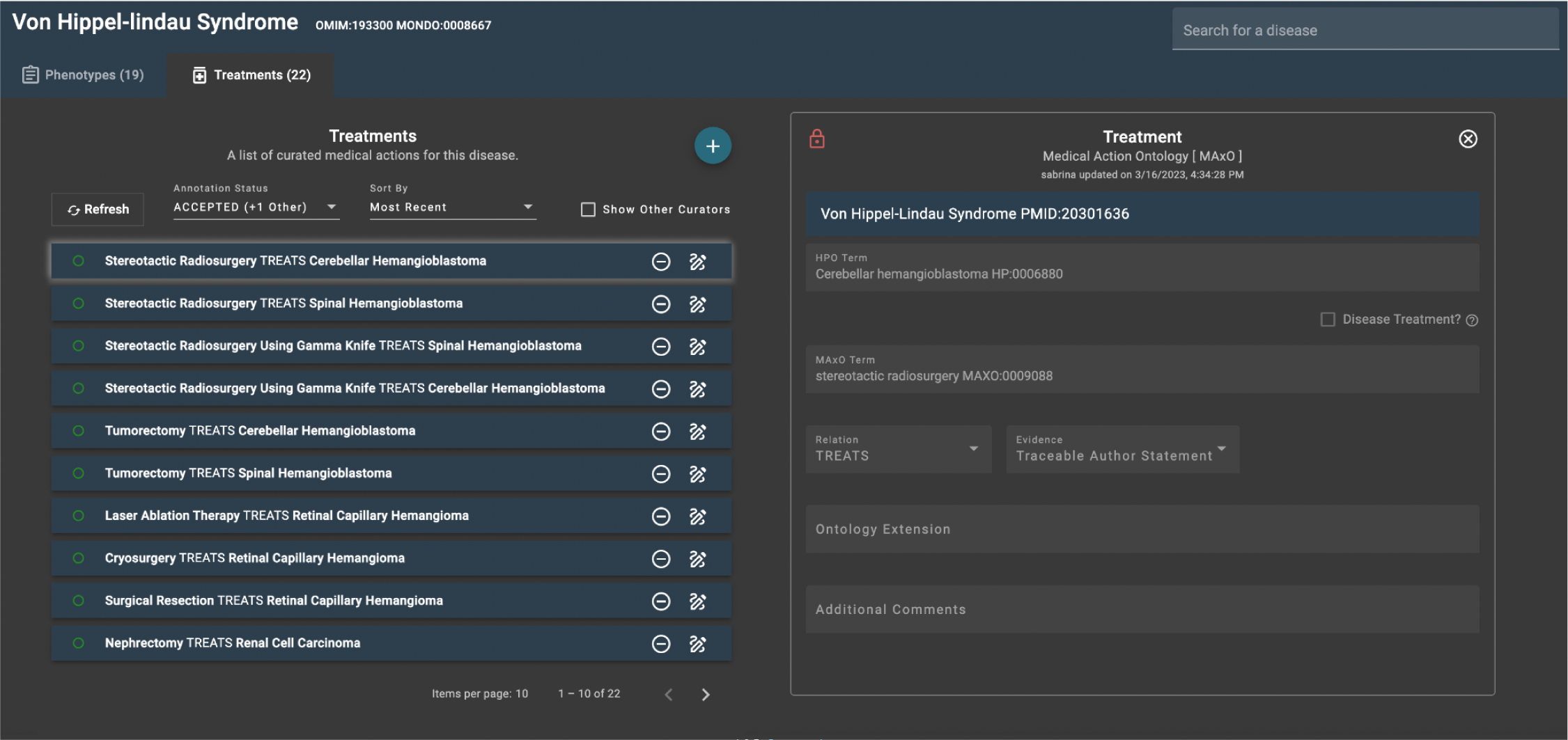
POET annotation page. The left side of the figure shows 10 MAxO annotations for Von Hippel-Lindau syndrome. On the right side of the figure, the annotation *stereotactic radiosurgery* (MAXO:0009088) has been entered, specifying that it treats Cerebellar hemangioblastoma (HP:0006880), which is a feature of the disease. Each annotation requires a source (in this case, PMID:20301636).^38^

## Community Involvement and POET

The Monarch curation team creates MAxO-HPO annotations using the POET web application; each annotation is created by one team member and vetted by at least one member who is a licensed physician. We developed POET to enable subject matter experts to participate and add more specialized, nuanced responses in their particular areas of expertise, regardless of their level of knowledge about building and using ontologies. We invite the community to participate in the annotation process. Registration in the POET system is free and a detailed tutorial is provided online. Participants are required to register with their ORCID identifier so that annotations can be attributed unambiguously.

## Discussion

The Medical Action Ontology (MAxO) aims to provide a computational representation of treatments and other actions taken for the clinical management of patients. The development of MAxO is closely coupled to the Mondo Disease Ontology (Mondo) and the Human Phenotype Ontology (HPO) and expands the scope of our computational modeling of diseases and phenotypic features to include diagnostic and therapeutic actions.

We envisage several key use cases for MAxO. Some rare-disease decision support software such as Phenomizer^39^ can suggest HPO terms of relevance to the differential diagnosis; given the current set of HPO terms and the current list of diseases deemed to be the most likely candidate, Phenomizer suggests HPO terms that, if present, would most strongly differentiate one or more of the candidate diagnoses from others.^39^ Using the diagnostic MAxO annotations presented here, clinicians can then assess whether a given investigation (e.g., *MRI of the abdomen*; MAXO:0000426) or referral to a specialist (e.g., *neurologist evaluation*; MAXO:0000716) would be the next diagnostic step most likely to elucidate the underlying diagnosis in a patient. HPO terms that characterize the diseases in a differential diagnosis can be grouped according to the diagnostic procedures that can investigate the presence of the abnormality represented by the term. For instance, three of the four HPO terms shown in Figure 3 can be ascertained by brain CT, while all four terms can be evaluated by MRI of the brain. If all four terms are important for the differential diagnosis, then it may be more appropriate to perform MRI as compared to CT.

The diagnostic odyssey that many individuals with RDs experience has been described many times in the literature.^12^ We are unaware of analogous studies concerning delays in the identification of appropriate treatment after an RD diagnosis is made, but the experience of the last author suggests that this is also a not infrequent problem. One difficulty is that many of the treatments available for RD do not treat the underlying disease but rather one or more specific manifestations of the RD. In some cases, the recommended treatment for the manifestation can differ from the treatment recommended for the same manifestation in other contexts. For instance, many monogenic epilepsies benefit from targeted treatments such as *pyridoxine supplementation* (MAXO:0001131) for individuals with Pyridoxamine 5’-phosphate oxidase deficiency.^40,41^ PubMed searches are not explicitly designed to find articles that discuss treatments of specific manifestations in specific diseases, and the anecdotal experience of the authors is that it can occasionally be challenging to find corresponding publications. MAxO is being used to annotate RD treatments by the Monarch Initiative and annotations will become available on the HPO website.

An increasing number of patients are being enrolled in RD clinical studies, and more and more therapeutics for RD are being approved.^10^ Moreover, non-pharmacological standards of care are especially important in RD management, as disease manifestations may include numerous aspects for which the primary treatment is not a drug, such as annual cardiac surveillance and pacemaker or implantable cardioverter-defibrillator implantation for cardiac management of myotonic dystrophy type 1 or the use of mechanical airway clearance devices in cystic fibrosis.^42,43^ Nutritional treatments, comprising medical foods or supplements and the avoidance of specific foods, are essential to the management of inborn errors of metabolism.^44^ Correspondingly, there is a need to make information about RD clinical management easily findable for clinicians and researchers. Clinical guidelines exist for some more prevalent RDs such as cystic fibrosis,^45^ and systematic literature reviews are available for some RDs developed by the Treatabolome group.^46^ Moreover, *GeneReviews^®^*has 864 chapters for RD or groupings of RD. These chapters provide resources and comprehensive information about the RDs and extensive information about the treatment and management of those diseases.^17^ MAxO could be used to make information in such resources more easily findable. Additionally, there are over 10,000 rare diseases^47^ and for many RDs, the only published information is in the form of case reports on small numbers of affected individuals. An added complexity for genetic diseases is that some therapies are only appropriate for the treatment of a specific genetic defect and may be ineffective or even harmful in individuals with the same disease phenotype but a different genetic cause,^48^ giving the need to unambiguously link a specific disease to a specific treatment even greater urgency, both to ensure timely treatment for amenable patients and to avoid harm as a result of giving the wrong treatment. There is a need to make this information about therapeutics and clinical management available to clinicians alongside diagnostic reports.

A range of variant and gene prioritization tools are used to prioritize candidate genes and diseases in clinical exome and genome sequencing.^49^ If there is an effective treatment available for one of the diseases in the list of candidates deemed to be most plausible, then diagnosticians should make every effort to make a definitive diagnosis so as not to miss an opportunity for treatment. MAxO disease-level annotations can be used to annotate diseases in software to support this goal.

To our knowledge, MAxO is currently the only ontology to specifically cover treatments and other medical actions. However, content related to medical actions is available in other ontologies and terminologies. For instance, SNOMED CT covers a wide range of healthcare domains beyond medical actions, including anatomy, clinical findings, medications, and devices,^50^ and the Thesaurus of the National Cancer Institute (NCIt) has a broad range of primarily cancer-focused terms including some for medical actions. Several other ontologies include some terms for medical actions, including the Ontology for Biomedical Investigations (OBI), Medical Subject Headings (MeSH), and the Ontology for General Medical Science (OGMS).^50–53^ Where appropriate, MAxO has included definitions from NCIt and provide cross references. However, none of these ontologies was developed to model medical diagnostics, treatments, and other actions, and thus not surprisingly, none of these ontologies has the depth of coverage of medical actions of MAxO. MAxO was designed to be used in tandem with HPO and MONDO and thus will be able to add value to many existing computational tools that use HPO or MONDO for bioinformatic research or clinical decision support systems.^13,54–57^ MAxO is made available under a Creative Commons Attribution 4.0, which will facilitate making data about medical actions findable, accessible, interoperable, and reusable (FAIR).^58^ Many OBO Foundry ontologies, such as MONDO, HPO, and OBI, have been constructed using logical definitions composed in the description logic of the Web Ontology Language (OWL).^16,59^ MAxO was developed using DOSDPs to generate terms and logical definitions with references to a total of 15 external ontologies. These definitions are resources that can be used as features for computational applications including knowledge graphs, searches, and machine learning.

## Outlook

MAxO was designed to promote standards for medical actions for use in numerous settings. The primary use case for MAxO is to provide a computable representation of information about treatments and other medical actions. The MAxO project includes a growing collection of annotations about diagnostic modalities that can be used to ascertain phenotypic abnormalities represented by HPO terms as well as annotations about medical actions for rare diseases. These annotations complement existing data in the HPO and Mondo projects and will contribute to more comprehensive computational models of disease.

MAxO provides a relatively high-level representation of medical actions, and does not in itself record dosages or other details of medical actions. Instead, we envision MAxO as enabling searches, machine learning algorithms, and clinical decisions support systems by providing an ontological representation of concepts related to medical actions and linking these to a peer-reviewed source for the clinician to consider. Currently, POET only supports therapy-related annotations, and suggestions for new diagnostic annotations need to be made via the GitHub issue tracker.

We anticipate that MAxO will be useful for many rare-disease data and knowledge resources, including resources intended to support treatment guidelines such as the Treatabolome.^46,48,60–64^ MAxO can be used to support the representation of medical actions in the Global Alliance for Genomics and Health (GA4GH) Phenopacket Schema.^65^ Additional future applications include support for decision support systems. MAxO will be developed alongside the HPO and MONDO to represent comprehensive models of rare disease. MAxO terms are applicable to any disease, and future work may include modeling of common disease clinical management.

## Funding

National Institutes of Health (NIH): NHGRI 1U24HG011449-01A1 and NHGRI 5RM1HG010860-04.

## Availability

MAxO is available at GitHub (https://github.com/monarch-initiative/MAxO) under a Creative Commons Attribution 4.0 International license. MAxO annotations are available on GitHub (https://github.com/monarch-initiative/maxo-annotations) under the same license.

POET is available at https://poet.jax.org/.

## Data Availability

All data produced are available online at https://github.com/monarch-initiative/MAxO and https://github.com/monarch-initiative/maxo-annotations

https://monarch-initiative.github.io/maxo-annotations/

## Notes

### Competing Interest Statement

The authors have declared no competing interest.

### Funding Statement

This study was funded by the National Institutes of Health (NIH): NHGRI 1U24HG011449-01A1 and NHGRI 5RM1HG010860-04.

